# A Quantitative and Narrative Evaluation of Goodman and Gilman’s Pharmacological Basis of Therapeutics

**DOI:** 10.1101/19007385

**Authors:** Brian J. Piper, Alexandria A. Alinea, John R Wroblewski, Sara M. Graham, Daniel Y. Chung, Livia R.M. McCutcheon, Melissa A Birkett, Steven S. Kheloussi, Vicky M. Shah, Qais K. Zalim, John A. Arnott, William A. McLaughlin, Pamela A. Lucchessi, Kimberly A. Miller, Gabi N. Waite, Michael Bordonaro

**Author notes:** Designated authors contributed equally (Livia R.M. McCutcheon, Melissa A Birkett, Steven S. Kheloussi, Vicky M. Shah, Qais K. Zalim, John A. Arnott, William A. McLaughlin, Pamela A. Lucchessi, Kimberly A. Miller, Gabi N. Waite, Michael Bordonaro).

## Abstract

**Objective:** Goodman and Gilman’s *The Pharmacological Basis of Therapeutics* (_GG_PBT) has been a cornerstone in the education of pharmacists, physicians, and pharmacologists for decades. The objectives of this report were to describe and evaluate the 13^th^ edition of _GG_PBT including: 1) author characteristics; 2) recency of citations; 3) conflict of interest (CoI) disclosure, and 4) expert evaluation of chapters.

**Methods:** Contributors’ (N = 115) sex, professional degrees, and presence of undisclosed potential CoI as reported by the Center for Medicare and Medicaid’s Open Payments (2013 to 2017) were examined. Year of publication of citations were extracted relative to comparison textbooks (Katzung’s *Basic and Clinical Pharmacology* (_Kat_BCP), and DiPiro’s *Pharmacotherapy: A Pathophysiologic Approach* (_DiP_PAPA). Content experts in pharmacy and pharmacology education provided chapter reviews.

**Results:** The percent of _GG_PBT contributors that were female (20.9%) was equivalent to those in _Kat_BCP (17.0%). Citations in _GG_PBT (11.5 ± 0.2 years) were significantly older than those in _Kat_BCP (10.4 ± 0.2) and _DiP_PAPA (9.1 ± 0.1, *p* < .0001). Contributors to _GG_PBT received three million in undisclosed remuneration from pharmaceutical companies (Maximum author = $743,718). In contrast, _DiP_PAPA made CoI information available. However, self-reported disclosures were not uniformly congruent with Open Payments reported data. Reviewers noted several strengths but also some areas for improvement.

**Conclusion:** _GG_PBT will continue to be an important component of the biomedical curriculum. Areas of improvement include more diverse authorship, improved conflict of interest transparency, and greater inclusion of more recent citations.

## INTRODUCTION

Educator and pharmacologist Louis S. Goodman (1906 – 2000) completed his undergraduate degree at Reed College and his MD at the University of Oregon Medical School in Portland, Oregon. Goodman and Alfred Gilman, his colleague and collaborator on nitrogen mustard investigations, published the first edition of *Pharmacological Basis of Therapeutics* (_GG_PBT) in 1941. Louis’s son Alfred G (Goodman) Gilman (1941-2015) received a Nobel prize for his work on signal transduction and served in various editorial capacities for the 5^th^ to 10^th^ editions. The first reviewer was “delirious in his appraisal of the book” and anticipated it would become the standard text in pharmacology.^1^ The eighteen hundred page 2nd edition published in 1956 was referred to as encyclopedic and indispensable.^2^ A reviewer noted that “all other related books seem to pale by comparison”.^3^ An evaluation of the 6th edition published in 1980 commended the extensive bibliography but was more measured and noted that although “this book is recommended to all those who prescribe drugs”, _GG_PBT had become “too large to be used by medical students as a routine textbook”.^4^ Hastings and Long referred to _GG_PBT as the “blue bible of pharmacology” and the “gold standard”.^5^ Rosenberg warmly commended this reference for dentistry and anesthesiology.^6^ However, despite the book’s esteemed and authoritative status, a subsequent reviewer alluded to one chapter where the text had “barely kept up” with the rapid pace of new therapeutic developments. Casavant also noted the omission of twenty of twenty-six newly approved medications in the 10^th^ edition^7^. The paucity of female authors and absence of potential conflict of interest (CoI) disclosure were concerns expressed about the 12^th^ edition published in 2012.^8^

An under-representation of females as authors has been identified in different fields and types of publications. Female first authorship in six, high-impact, general medical journals increased from 27% in 1994 to 37% in 2014.^9^ Senior authorship by women in cardiology journals doubled from 6% in 1996 to 12% in 2016.^10^ Textbook authorship, which often involves large teams, is highly variable and ranged from half (53%) female for an advanced pharmacy textbook, the 2014 9^th^ edition of DiPiro’s *Pharmacotherapy: A Pathophysiological Approach* (_Dip_PAPA),^11^ to one out of seven (14%) in Yagiela’s *Pharmacology and Therapeutics for Dentistry*.^8,12^

Conflicts of interest (CoIs) were defined as “a set of circumstances that creates a risk that professional judgment or actions regarding a primary interest will be unduly influenced by a secondary interest”.^13^ The transparency of conflicts of interest has become increasingly ubiquitous for primary sources^14^ including clinical trials,^15^ undergraduate medical education,^16^ continuing medical education,^17^ point of care computerized sources,^18^ meta-analyses,^19^ and clinical practice guidelines.^20^ The US Physician Payments Sunshine Act of 2010 required that all compensation (≥ $10) to doctors of medicine, osteopathy, dentistry, dental surgery, podiatry, optometry, and chiropractic medicine (i.e. PharmDs, PhDs, PAs, and NPs were not covered although this subsequently changed for PAs and NPs) by manufacturers of drugs and medical devices be reported to the Centers for Medicare and Medicaid Services (CMS) and made available on its public website. Disclosure of conflicts of interest was provided in the preface to a psychopharmacology textbook^21^ and in DiPiro’s *Pharmacotherapy: A Pathophysiological Approach* but this practice is currently uncommon. The database, ProPublica’s Dollars for Docs (PDD), originally covered only fifteen pharmaceutical companies. Using the first generation of this database, we found that authors and editors of four biomedical textbooks had received $2.4 million, primarily for speaking and consulting, which was undisclosed to readers. One-quarter of the contributors to _GG_PBT, 12^th^ edition, had an undisclosed patent.^8^

Here, we extend upon earlier educational research^8, 12^ by assessing the most recent (2018, 13^th^ edition) of _GG_PBT. This includes quantitative descriptive measures of author characteristics (education, sex), a citation analysis relative to other pharmacology (_Kat_BCP) and pharmacotherapy (_DiP_PAPA) textbooks and characterizing the presence of potential CoI using more comprehensive databases. Content experts also provided detailed assessments in their specialty areas.

## METHODS

### Procedures

The contributors page of the 13^th^ edition of Goodman and Gilman’s *The Pharmacological Basis of Therapeutics* (_GG_PBT) ^22^ was consulted to obtain professional degrees of authors and editors (N = 115). Sex was determined by a Google search or consulting the National Provider Identifier (https://www.cms.gov/Regulations-and-Guidance/Administrative-Simplification/NationalProvIdentStand/). The presence of CoI was deemed exempt by the Wright Center Institutional Review Board and evaluated with two publically available databases, the Center for Medicare and Medicaid Service’s Open Payments (_CMS_OP) and ProPublica’s Dollars for Docs (_PP_DD). These databases differ slightly with respect to when potential CoI information is made available and to details (e.g. specific companies and products) about disclosures, with ProPublica’s Dollars for Docs being updated less frequently but providing greater depth. Payments were obtained for 2013 to 2017. The second generation of _PP_DD reported on $9.2 billion in payments from two-thousand companies to nine-hundred thousand physicians. Among _GG_PBT authors with a US affiliation (N = 109), 42.3% had professional degrees that were covered by the Sunshine Act (i.e. PhDs and PharmDs were not covered). Year of publication of each reference for 72 chapters including the Appendix (N = 3,576) was obtained. The publication year of all citations of Katzung’s *Basic and Clinical Pharmacology* (_Kat_BCP, 2018, N = 1,777)^23^ and _Dip_PAPA (9^th^ ed., 2017, N = 13,389)^11^ including eChapters were also obtained for comparison.

For the narrative reviews, content experts were consulted. Experts included academic pharmacists, physiologists, molecular biologists, a psychiatry resident, a neuroscientist, a neurologist, and a bioinformaticist. Experts were instructed to succinctly provide strengths and limitations and, if applicable, to make comparisons with other textbooks they use. Some provided assessments of a specific chapter, others provided an evaluation of an entire section. Two recent graduates from a Master’s in Biomedical Sciences program provided student perspectives. The goal was not to be comprehensive of all 71 chapters but instead to provide at least a sampling from most sections.

### Data-analysis

Statistical analysis was completed with Systat, version 13.1 with *p* < .05 considered significant. Figures were prepared with GraphPad Prism. Sex of contributors of _GG_PBT was compared to other pharmacy and biomedical textbooks obtained previously.^8, 11^ Age of references was calculated as the difference between publication year and the copyright year. The similarity between CoI databases was evaluated with a Pearson correlation and t-test. Variability was expressed as the SEM with a *p* < .05 considered statistically significant.

## RESULTS

### 1. Quantitative

In Goodman and Gilman’s *The Pharmacological Basis of Therapeutics* (_GG_PBT), there were 115 authors, of which 32 contributed to more than one chapter. The highest degree was MD for half of authors (47.0%), PhD for two-fifths (45.2%), and the remainder (7.8%) PharmDs. The preponderance (90.4%) of authors were based in the US.

One-fifth (20.9%) of authors were female or 19.0% if including multiple chapter contributions.

Female authorship in _GG_PBT was equivalent to that of Katzung’s *Basic and Clinical Pharmacology* (_Kat_BCP, 17.0%, *p* = .52) but less than Remington’s *Science and Practice of Pharmacy* (37.0%, χ^2^(1) = 8.84, *p* < .003) and Koda-Kimble and Young’s *Applied Therapeutics* (53.0%, χ^2^(1) = 8.84, *p* < .001).

Figure 1A shows that references in _GG_PBT were 12.3% older (11.5 years) than those in _TK_BCP (10.4, *t*(5,321) = 4.42, *p* < .0005). The DiPiro’s *Pharmacotherapy: A Pathophysiological Approach* (_DiP_PAPA) citations (9.1 years) were also more recent than _GG_PBT (*t*(16,951) = 15.59, *p* < .0001). Similarly, additional analysis determined that more references were > a decade old for _GG_PBT (49.1%) compared to _Kat_BCP (44.7%, χ^2^(1) = 9.20, *p* < .002) or _DiP_PAPA (34.0%, χ^2^(1) = 278.21, *p* < .0001). _Kat_BCP also differed from _DiP_PAPA on this measure (χ^2^(1) = 78.44, *p* < .0001). Figure 1B depicts reference age by section of _GG_PBT. The Drugs Affecting Gastrointestinal Function were significantly newer than all other sections. Conversely, Neuropharmacology references were 6.7 years older than Gastrointestinal and also significantly less recent than all other sections. There were some broad similarities between _GG_PBT and _Kat_BCP (Figure 1C) with oncology citations being more recent, endocrinology intermediate, and neuroscience the oldest. The gastrointestinal, as well as gynecological, citations were most recent relative to many other sections in _DiP_PAPA (Figure 1D).

**Figure 1.**
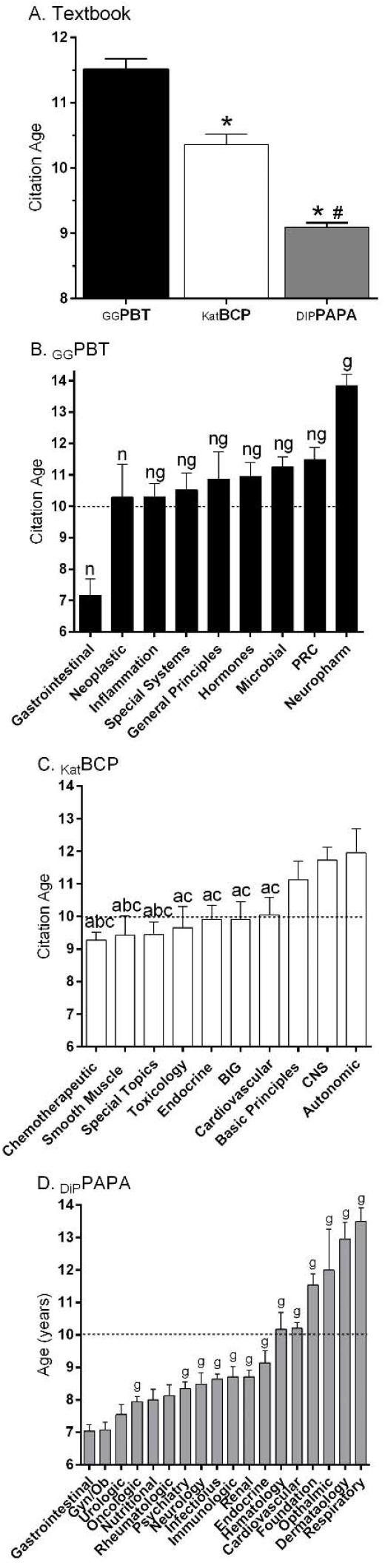
Citation age (± SEM) for *Goodman and Gilman’s Pharmacological Basis of Therapeutics* (_GG_PBT, 13^th^ edition, 2018) relative to *Katzung’s Basic and Clinical Pharmacology* (_Kat_BCP, 14^th^ edition, 2018) and *DiPiro’s Pharmacotherapy: A Pathophysiologic Approach* (_DiP_PAPA, 10^th^ edition, 2017, A. ^*^*p* < .01 versus _GG_PBT, ^#^*p* < .0005 versus _Kat_BCP). Age by section of _GG_PBT (B, ^n^*p* < .0005 versus Neuropharmacology; ^g^*p* < .0005 versus Gastrointestinal), _Kat_BCP (C, ^a^*p* < .05 versus Autonomic, ^b^*p* < .05 versus Basic Principles, ^c^*p* < .05 versus Central Nervous System (CNS). PRC: Pulmonary, Renal, Cardiovascular; BIG: Blood, Inflammation, Gout) and _DiP_PAPA (D, *p* < .01 versus Gastrointestinal Disorders, and Gyncologic and Obstetric Disorders).

The similarity of the conflict of interest databases ProPublica’s Dollars for Docs (_PP_DD) and Medicaid Service’s Open Payments (_CMS_OP) was examined with two complementary analyses. The correlation between the total received from 2013 to 2016 (i.e. the most recent available in both databases) was high (*r*(24) = +0.999, *p* < .0005; Supplementary Figure 1). However, the mean amount received was eighteen-hundred dollars higher for _CMS_OP ($82,923 + 24,404) than _PP_DD ($81,105 ± 24,122), a non-significant difference (*t*(25) = 1.59, *p* = .13).

Total undisclosed remuneration received by twenty-seven _GG_PBT authors (57.4% of eligible US based physician-authors, 96.3% male, Minimum = $23, Maximum = $743,718) from 2013 to 2017 was $2.97 million of which almost three-quarters (71.4%) went to the top five authors. The most compensated author received $493,536 (66.0% of their total) for royalty or licenses and $250,108 (34.0%) for ownership or investment interest. One of the dermatological pharmacology authors received $493,536 for ivermectin, a head lice treatment. The psychosis and mania chapter author received $238,413 for payments related to two atypical antipsychotics. The oncologist author of “Hormones and Related Agents in the Therapy of Cancer” received $228,493 for five breast cancer agents.

Additional analyses were completed on _DiP_PAPA which provides CoI information. Approximately one out of every twelve contributors (20/233 or 8.6%) self-reported a potential CoI. Among the 35 eligible (MD or DO with a US affiliation) authors, 74.3% had a _PP_DD entry (Min = $14, Max = $729,695, total = $1.8 million). However, some discrepancies were identified. A status epilepticus author whose disclosures were self-reported as “none” received a half-million for anti-epilepsy drugs according to _PP_DD. Two authors who did not report disclosures to _DiP_PAPA accepted $70-350 thousand in potentially relevant remuneration from 2013 to 2016 (Supplemental Table 1).

### 2. Narrative Reviews

Open-ended evaluations were provided on sixteen chapters including representation of seven of the nine sections of Goodman and Gilman’s *The Pharmacological Basis of Therapeutics* (_GG_PBT).

#### 2.1 General Principles

Chapter 2, “Pharmacokinetics: The Dynamics of Drug Absorption, Distribution, Metabolism, and Elimination” is a comprehensive and thorough explanation and discussion of the basic concepts of pharmacokinetics. The characteristics and processes of absorption, distribution, metabolism, and elimination are reviewed respectively, including an in-depth discussion of the types of transport across membranes and the influence of pH. The first-pass effect as well as the benefits and limitations of various routes of drug administration are also explored. Potentially beneficial additions to the discussion are further examples of specific drugs and their routes of administration, as well as the factors that influence onset of action. There is a brief discussion of rectal administration that could be improved by expanding upon the reasoning behind its incomplete and irregular absorption. Distribution is briefly explored, including a discussion of the differences of distribution among various types of tissues as well as an explanation of protein and tissue binding. While the bone portion of this section focuses mostly on tetracycline antibiotics, an introduction to the pathophysiologic changes in bone that make distribution difficult overall would be helpful. The next section on metabolism is a simplistic but comprehensive explanation of phase one and two reactions, as well as first and zero-order kinetics. A potentially beneficial supplement to this section could be to include examples of when products of metabolism make the drug more toxic (eg. acetaminophen toxicity). The excretion section focuses primarily on renal excretion but also includes that by other routes. Learning is aided by highly beneficial figures that depict renal drug handling and a helpful visual of the afferent and efferent arterioles. A recommendation for improvement is to move the discussion of drugs in breast milk to the distribution section. While placental transfer is discussed in the distribution section, breast milk distribution and potential for ion trapping is not. Tissue distribution is more relevant than elimination when discussing drugs in breast milk. The final section discusses clinical pharmacokinetics and the “four most important parameters governing drug disposition.” While this section is extensive and includes strong drug examples to explain clinical pharmacokinetics, it could be confusing to the reader that components of “clinical pharmacokinetics” are separated from ADME. It may be beneficial to integrate this section into earlier content. Some examples include moving the discussion of extent and rate of absorption under absorption, clearance under excretion, and volume of distribution under distribution. There is a helpful explanation of maintenance and loading doses, therapeutic window, and dosing intervals along with drug examples with each. However, the explanation of therapeutic drug monitoring is somewhat limited and potentially too generalized. Additional examples of therapeutic drug monitoring (for example, a comparison of therapeutic drug monitoring used for vancomycin, aminoglycosides, and warfarin) would be beneficial to the reader in order to understand the parameters that influence different timing and results of monitoring.

Chapter 3, “Pharmacodynamics: The Molecular Mechanisms of Drug Action” includes a comprehensive and highly detailed discussion of three primary areas of pharmacodynamics including basic concepts, mechanisms of drug action, and signaling pathways. The first section provides a detailed explanation of different receptor types as well as a short discussion of the increased development of biologic agents. The definitions of agonists, antagonists, and their subtypes are provided. A useful addition to this section would be drug examples of agonists, antagonists, and their subtypes in order to illustrate these concepts further. Specificity of drug receptors and tachyphylaxis are explored, followed by a detailed explanation of affinity, efficacy, and potency. Graphs and mathematical models are used to elaborate upon these concepts. Individual and population pharmacodynamics as well as factors that affect the variability of drug dosing are included. There is also a useful section that expands on the individual patient characteristics that contribute to variability of dosing. There is a section on drug interactions, but it does not mention the effect of CYP enzymes on drug interactions. While this is typically categorized in pharmacokinetics, it may be beneficial to mention their substantial effects on drug levels here. The following section explores the mechanisms of drug action, and expands upon the effects of drugs on ligands, extracellular responses, intracellular pathways, and ions. The mechanisms of anti-infective drugs are also reviewed, which can potentially be eliminated in the overall discussion, or converted to an example instead. There is a helpful review of the structure and function of specific receptor types including second messenger systems and other signaling pathways. Tachyphylaxis and desensitization are discussed again here and could be more beneficial if the discussions on this topic were condensed to one area of the chapter. The chapter ends with an extensively detailed exploration of diseases associated with transcription and translation along with the pharmacotherapies that treat them. Considering that this is an introductory chapter on pharmacodynamics, it may be more useful for this information to be included at a later point in the book. It may be advantageous to consider narrowing this topic to only basic pharmacodynamic concepts.

The General Principles section further includes Chapter 7, “Pharmacogenetics”, which provides an overview of the terminology and principles that describe the genetic variations that contribute to drug response phenotypes. A description of the applications of individual genotyping data to inform clinical decisions is provided. Different types of genetic variations which can affect drug response phenotypes are outlined. Each type of genetic variation is linked to a description regarding a change in biological function, which in turn is linked to the phenotype. Emphasis is thereby placed on interpreting the genetic variations in terms of the anticipated functional consequences.

As a possible improvement, a further systematic categorization of the linkages between genetic variations and drug response phenotypes may be provided. Consider the categorizations of the linkages given by Katsila et al. ^33^ The linkages are described in terms of the overall genetic category (poor metabolizer, extensive metabolizer, therapeutic target etc.), the functional effect, the possible phenotype associated with an active drug, and the possible phenotype associated if a pro-drug. Such categorization may help the reader to gain better understanding of apparent themes in the drug response phenotypes. For example, regarding prodrugs, the reader may further expect that an adverse effect would occur in the presence of the drug-activating enzyme that has a relatively high activity.

Chapter 7 also advances an understanding of the functional consequences of genetic variations in terms of complete or partial loss of function. A discussion of the increased relative effects of variations at positions with different levels of evolutionary conservation is provided. But in the previous (12^th^) edition of *Goodman and Gilman’s Pharmacological Basis of Therapeutics* (_GG_PBT), there was greater delineation of the descriptions of evolutionary conservation versus chemical conservation. It may be helpful to reintroduce that back into the chapter to clarify these important concepts. It may then become clearer that positions in a protein which are highly evolutionary conserved may not allow for an amino acid which is chemically similar if that substituted residue does not enable the protein’s specific function.^34^ In the previous edition, it was more evident that one would need to consider evolutionary conservation and chemical similarity in a somewhat separate light.

As a welcome addition to the current edition, there was an informative discussion of the need to consider common versus rare variations of a population when assessing the effects on drug response phenotypes. Discussions are provided regarding how studies are designed to detect common variations and rare variations using candidate gene association studies and genome-wide association studies. It is made apparent that both rare and common variations will need to be considered when predicting the drug response phenotypes. The changes from the 12th edition to the 13th edition with regards to the inclusion of the effects of rare and common variants appear to be due in part to changes the contributing authors. For the 12th edition, the authors were Mary V. Relling and Kathleen Giancomini, while the author for the 13th edition is Dan Roden, whose research focus is on characterizing such variations.

The number and breadth of the described genetic variations provide the reader with an appreciation how pharmacogenomic information will be used to predict drug response phenotypes and to inform treatment strategies. Consider Table 7-2, entitled “Examples of Genetic Polymorphisms Influencing Drug Response”. The presentation of the example drugs provides the reader incentives to understand the related topics which are presented throughout the remainder of the textbook. To aid with that, hyperlinks may be added to Table 7-2 that connect to these topics.

#### 2.2 Neuropharmacology

Chapter 15, Drug Therapy of Depression and Anxiety Disorders, includes several strengths. Figure 15.1 illustrates detailed information about the variety of antidepressant mechanisms or action. It provides a list of long term cellular regulatory changes in addition to increasing neurotransmitter “dwell time” in the synapse. The development of ketamine and other approaches as novel antidepressants are well described. Information about pharmacokinetics and CYP action is helpful in a practical way to predict, and avoid, drug interaction. This chapter provides a general summary of the basic treatments for anxiety and depression.

As a weakness, chapter 15 does not place very much emphasis on the dopamine or histamine systems/actions in antidepressant effects or depressive etiology. Information about side effects, therapeutic lab, wash-out periods, serotonin syndrome and discontinuation effects are dispersed throughout the text and are not highlighted or emphasized as key pieces of information in designing a treatment strategy. At the beginning of the chapter, depression is immediately categorized into bipolar I and II. It might be more accurate to introduce mood disorders more generally, with an emphasis on depressive symptoms/diagnoses. The authors suggest that there has been limited progress in developing animal models sensitive to antidepressants and anxiolytics. Anxiety is described as “a normal human emotion that serves an adaptive function,” however depression is not.

Another concern is that there are places where information is presented without research citations, such as “Anxious patients appear to be particularly prone to severe discontinuation reactions with certain medications such as venlafaxine and paroxetine; therefore, slow-tapering is required.” The chapter does not include information about options for drug-treatment-resistant symptoms or suggestions for combination approaches.^21^ As this is a pharmacology text, that is reasonable. However, it might be helpful to have at least mention of electroconvulsive therapy or transcranial magnetic stimulation, so readers are better informed of other options if drug treatment has been unsuccessful. More specialized textbooks^21^ provide at least a nod that the nomenclature for the “Selective” Serotonin Reuptake Uptake Inhibitors is an inaccurate oversimplification.^24^

Chapter 20, “Opioids, analgesia, and pain management” is well organized and includes an emphasis on history, receptor signaling, the pathophysiology of pain, tolerance, withdrawal, and medical chemistry for this important drug class. On rare occasions, there is some unusual content and minor areas for reconsideration. Mentioning that opioids do not bind to sigma receptors, twice, may be less useful for new members of the field. The space devoted to specific opioids does not show any simple relationship with their current use patterns.^25^ Meperidine and normeperidine, despite concerns with CNS excitation and seizures, are mentioned four times more commonly than the ubiquitous hydrocodone. Tramadol is described, twice, as a “weak opiate agonist”. Katzung’s *Basic and Clinical Pharmacology* (_Kat_BCP) and others have a more nuanced description of the mechanism of action of this agent and the importance of desmethyltramadol.^26^ The term “addict” or “addicts” are used eight times versus zero for the less stigmatizing “opioid use disorder”.^27^

Chapter 23 “Ethanol” begins with a brief description about the historical use of alcohol in human civilization as well as epidemiologic overview of problems associated with it; this is quite helpful. There also is a practical overview about alcohol content of different beverages as well as information about estimating the blood ethanol concentration in end expiratory alveolar air. Excellent sections with pharmacological properties of methanol and ethanol and its effects are arranged by system. The addition of a Shakespearean quote added emphasis and served as a nice anecdote to illustrate the overview. This is also a good overview about the postulated neurological pathways that are thought to be involved in tolerance and dependence. Descriptions of teratogenicity, genetics, and drug interactions are adequate. Information about treatment of alcohol withdrawal is lacking. No information about choice of drug therapy for withdrawal as well as symptoms triggered therapy protocols (the Clinical Institute Withdrawal Assessment or CIWA protocol for example) for assessment and treatment is provided. This is a surprising omission as alcohol withdrawal is very prevalent and is associated with considerably increased mortality and morbidity as compared to opioid withdrawal.

#### 2.3 Modulation of pulmonary, renal, and cardiovascular function

Chapter 25 has the challenging task of introducing normal renal structure and function as a basis for the mechanistic understanding of drugs that affect the renal excretory function. The chapter does an incredible job in covering clinically relevant renal concepts. However, it is somewhat arbitrary in what is introduced in the chapter’s introduction as “normal” and what is mentioned later, because it is necessary to mention to understand the mechanics of diuretics and other drugs. Some figures and tables are very helpful (e.g., Table 25-1), while others are overloaded and do not add much value (e.g. Figure 25-2). Overall, it is a good chapter, despite the fact that every pathophysiologist will miss some mechanistic understanding of normal (e.g., Mg^2+^ reabsorption) or abnormal processes (e.g. reduction of urinary Ca^2+^ excretion by thiazide diuretics).

#### 2.4 Hormones and hormone antagonists

Chapter 48 “Agents Affecting Mineral Ion Homeostasis and Bone Turnover” provides a thorough review of pharmacological agents used in treating and preventing mineral ion imbalances and bone metabolism. The tables and figures are highly useful and summarize the main points of discussion, particularly the drug summary table at the end of the chapter, which provides a quick reference for understanding the pharmacological agents, their uses, and their clinical effects. This chapter will be useful for both the medical student and seasoned healthcare professional in that in addition to discussing all of the major pharmacological players involved in treating ion and bone diseases, it makes great attempts to review and contextualize these treatment approaches by providing significant background information. For example, this chapter contains a review of the basic hormonal control mechanisms involved in ion homeostasis, target organs/systems, relevant bone cell physiology, and summary of the biology of disorders of mineral homeostasis and bone. The chapter focuses on general concepts that have high relevance in the clinical setting and are useful for directly understanding the underlying pharmacology. This approach, while on the whole effective, does sometimes come at the expense of missing some detailed but relevant information. For example, the chapter really does not discuss osteoclast function in depth or bone resorption/remodeling effectively which would aid in the reader’s understanding of drugs like bisphosphonates. This chapter also contains a decent summary discussing the integrated approach to prevention and treatment of osteoporosis. This section does a more than adequate job of generally summarizing the field with respect to the treatment strategies used for managing and preventing osteoporosis.

#### 2.5 Gastrointestinal (GI) pharmacology

Chapters 49 through 51 focus on gastrointestinal (GI) disorders. There are several positive aspects to these chapters. First, the physiology and pathogenesis overview of each chapter and pharmacology of each medication class is brief yet manages to remain quite detailed. The use of figures throughout allows information to be more easily digested. Also, as a new text, this resource incorporates information on recently approved agents, including medications like eluxadoline (Viberzi^®^) and vedolizumab (Entyvio^®^), which naturally would be omitted by older resources. Finally, information on each drug and drug class appears complete and accurate. Even adverse events recently identified by newer studies, like dementia and chronic kidney disease for proton pump inhibitors (PPIs), are included.

On the other hand, there are several drawbacks as well. This section groups a large number of topics into a small number of chapters, which could make readings challenging to assign to students. For instance, to discuss a topic like *Helicobacter pylori* infection, one must first learn about PPIs early in Chapter 49, then navigate through about two-thirds of the chapter before finally coming across a brief discussion on managing the disorder.

The organization of Chapter 50, which covers motility disorders, emesis, and biliary and pancreatic disease, is particularly hard to follow. The chapter begins by introducing antimotility agents used in small populations and only available through limited access programs, if at all. Meanwhile, very common motility disorders like diarrhea and constipation are buried throughout the lengthy chapter. Also, the complications of cirrhosis, an important GI disorder, are mixed in only in short blurbs and in a confusing manner throughout the chapter. A more complete discussion is vital and its absence is a concerning flaw of this section.

Also, while it may be fitting to limit or omit information on medical interventions, such as fundoplication surgery, there is limited coverage on nonpharmacologic interventions, which are some of the most common and important recommendations pharmacists can make to patients. Additionally, some aspects of these chapters assume a baseline understanding, which may not be beneficial for students who do not possess this prior knowledge. For instance, terms like distal proctitis are left undefined, yet are used freely when describing the therapeutic role of certain agents. Finally, compared to other resources, this section uses fewer tables, making differences between medications or diseases more difficult to establish.

Overall, chapters 49 to 51 allow individuals to build a strong foundation in pharmacologic treatment of GI disorders by providing a great deal of information in a small number of chapters. However, this section is not without its flaws. Improvement is needed with regard to organization, coverage of cirrhosis, information on nonpharmacologic interventions, and the use of tables to present data.

#### 2.6 Chemotherapy of infectious diseases

Chapters 52, 56, 57 and 58 focus on infectious diseases. Chapter 52 is a general chapter focusing on the general principles of antibacterial therapy. There are a few positive aspects to this chapter including the step by step description of which antibiotic type (prophylactic, definitive, suppressive, etc.) should be utilized in the ID process and detailed explanations of each type. The overview of the pharmacokinetics of antibiotics is very detailed and the use of figures throughout the descriptions provides visual explanations to better understand the information. The explanation on medication resistance is very thorough and ensures that detailed information for resistance of each medication class does not have to be discussed in each respective chapter.

Chapter 52, general principles of antimicrobial therapy, was very informative but does have some disadvantages which make it somewhat difficult to follow. The pharmacokinetics section, though detailed and useful, covers information beyond the need of healthcare providers. The descriptions of E_max_ and K_a_, including equations, are explained well but are not necessary to fully describe the antibiotic classes. This information is not useful when determining what antibiotic to utilize, nor does it explain how dosing is affected based on these values. The explanation of time-dependent, concentration-dependent and antibiotics with post-antibiotic effect are explained very well; however, a comprehensive table detailing what medications fall into each category is not provided. Most other textbooks include a description of what an infection entails and appropriate selection of antibiotics; however, this chapter jumps directly into the pharmacokinetic information.

Chapters 56-58 discuss various antibacterial agents with descriptions of structure activity relationships, mechanism of actions, antibacterial spectrum, pharmacokinetics and indications for each medication within each respective class. The organization of the chapters has an excellent flow and is consistent from class to class. In chapter 56, the discussion on medications used for urinary tract infections is not consistent with the other medication classes; however, the information provided is very thorough and does incorporate the same information provided by the other class of medications, just in a different manner. Even though there are copious amounts of information provided on each respective class and each medication within the class, the format and explanations are done in a very lay method allowing non-healthcare providers to understand the material as well. The inclusion of historical facts about certain medication classes improves the reading quality and provides background information on either the class or the antibacterial spectrum of the class. In chapter 57, the inclusion of the carbenicillin class is necessary to explain the class as a whole. Other texts will exclude these medications and only discuss piperacillin/tazobactam; however, including the medications not available in the United States, such as ticarcillin and mezlocillin, increase the credibility of piperacillin. Most texts have difficulty explaining the cephalosporin generations; however, chapter 57 does an impeccable job detailing out medications in each generation and relating it back to the antibacterial spectrum. The summary table at the end of chapter 57 chapter 58 is all-inclusive and does a superb job at summarizing the chapter.

These chapters are very thorough and useful to a variety of healthcare providers; however, there are a few limitations within these three chapters which provides information that is not needed or lacks information in certain areas. In chapter 56, when discussing sulfonamides, several medications are referenced which are not commonly used, including one which is not approved for use in the United States. This excess information makes it more difficult to find the relevant information for medications more commonly utilized. One major limitation within these three chapters is the lack of tables throughout the chapter to simplify indications and dosing. All indications are separated into respective paragraphs which makes it more difficult to follow, providing tables after each class highlighting the indication and doses would allow readers to connect all medications within the class. It is understandable that not all new medications could be included due to the publication process being lengthy; however, four medications released within the last year are not included, two of which make a huge impact in practice. Delafloxacin (Baxdela^®^) and the combination of meropenem and vabobactam (Vabomere^®^) were introduced within the last year, but they have not warranted major changes; however, the combination of ceftazidime and avibactam (Avycaz^®^) and the combination of ceftolozane and tazobactam (Zerbaxa^®^), medications introduced specifically to fight off resistant pathogens such a carbapenemase-resistant enterobacteriaceae, are essential for healthcare providers to know about due to increased resistance to other medications in recent years.

Overall, these chapters allow healthcare providers to obtain the necessary pharmaceutics information needed to make decisions in patient care; however, improvements in formatting and the inclusion of newer medications would benefit the text as a whole.

#### 2.7 Pharmacotherapy of Neoplastic Disease

Chapter 65, “General Principles in the Pharmacotherapy of Cancer” is an excellent and reasonably comprehensive background to cancer pharmacology. Strengths of this chapter include: (a) distinguishing slower-growing cancers that have a smaller proportion of actively cycling tumor cells and are thus less responsive to drugs that target the cell cycle; (b) stressing the importance of combinatorial therapy, particularly combinations of molecularly targeted drugs and immunotherapy with more generalized cytotoxic chemotherapy; and (c) a discussion of the various challenges of molecular testing to determine those patients for whom specific targeted therapies would be most efficacious. With respect to the last issue, the chapter not only discusses the issue of tumor heterogeneity but also properly cites inherited genetic variation (and not just tumor-specific variation) as a factor affecting treatment response. One criticism is, in the section on resistance, it is stated that the resistant cells pre-exist the treatment which selects for these cells. While this no doubt occurs in the (vast) majority of cases (and the evidence for this for kinase inhibitors is discussed in Chapter 67), it should be noted that resistant cells can occur during treatment as a result of random mutations, unrelated to treatment, that may occur, particularly in tumors exhibiting a hypermutable condition (e.g., MSI+). Also, the much more controversial issue of adaptive mutation, which is starting to attract serious attention from a subset of researchers, could have at least been mentioned in passing. For example, there has been a report of the possible role played by adaptive mutation in the development of resistance to the androgen receptor antagonist bicalutamide in prostate cancer cells.^28^

Chapter 66 goes into great detail about several important classes of cytotoxic drugs, mostly those that block cell division and/or promote apoptosis, but also the differentiating agent all-trans retinoic acid (ATRA). Alkylating agents and platinum analogs come in for a particularly extensive review, and one that does justice not only to mechanisms of action and therapeutic efficacy, but also the (often serious) side effects of these agents. The chapter also makes an important distinction between bifunctional agents and monofunctional methylating agents, which is helpful.

Chapter 67, “Pathway-Targeted Therapies: Monoclonal Antibodies, Protein Kinases Inhibitors, and Various Small Molecules” delves into a detailed examination of pathway-targeted therapies, centered on small molecule inhibitors and monoclonal antibodies, two approaches that are contrasted with respect to range of action and to their side effects (the small molecules tend to have both a greater range of desirable activities as well as negative side effects). Similar to the examination of cytotoxic therapies in the preceding chapter and to the more general introduction of chapter 65, attention is paid to issues of mechanism of action, matching specific therapy to the appropriate type of cancer including genetic variation of the cancer, preexisting inherited genetic variation of the patient that affects response, side effects, and development of resistance. Also, combinatorial therapy is touched upon, an approach particularly useful with the monoclonal antibodies. The sections on angiogenesis inhibitors and immunotherapy (particularly the immune checkpoint inhibitors) was quite good, and this reviewer positively notes the discussion of combinatorial immune checkpoint therapy targeting both cytotoxic T lymphocyte–associated protein 4 (CTLA-4) and programmed death-1 (PD-1). There was some discussion about how to overcome resistance; for example, resistance to the MEK inhibitor trametinib can be overcome by combinatorial therapy with the BRAF inhibitor dabrafenib, and there is also discussion about dealing with imatinib resistance. This is all significant; if anything, more on this topic can be included; the same can be said about ameliorating side effects of these agents. There was some decent discussion on this latter topic; particularly useful was the note about colony stimulating factors used to deal with hematopoietic toxicity of many anti-cancer therapies. Certainly, an expanded analysis of such topics is always welcomed. The section on histone deacetylase inhibitors was adequate, but there could have been a discussion about butyrate, a fermentation product of dietary fiber, which is a histone deacetylase inhibitor and one that may significantly mediate the anti-cancer properties of dietary fiber for the colon. These topics could have been broached, which would have also allowed for a discussion of how naturally occurring agents in food may exert preventive action against cancer. Although the book is focusing on pharmacological therapy, the overlap between pharmacology and “medicinal food,” as well as that between prevention and treatment, could have been productively addressed for the sake of completeness. Chemoprevention is a legitimate topic in the field of cancer, both with respect to natural products as well as, perhaps, other pharmacological agents. One issue that could have been included as additional detailed discussion is the approach of inducing apoptosis through hyper-activation (rather than inhibition) of signaling pathways the cancer cell is “addicted to.” There also could have been some discussion of cutting-edge small molecules in clinical trial that target signaling pathways, such as ICG-001-like compounds that inhibit CBP-mediated Wnt signaling. Further, by connecting issues such as targeting signaling pathways, hyperactivating signaling pathways, and molecularly targeted drugs, one could cite the finding that some histone deacetylase inhibitors can hyperactivate Wnt signaling to induced apoptosis of colorectal cancer cells in culture, pointing to a possible novel therapeutic approach. That underscores one weakness of these chapters: while they tend to give an excellent overview of existing therapies, there is not much about possible novel future approaches.

Chapter 68 offers sound coverage on the role of hormone modulation for cancer therapeutics. Two main approaches were addressed. First, there was discussion about glucocorticoids, which can not only be used for anti-cancer treatment, but also to ameliorate side effects from other forms of cancer therapy. Second, hormone-based therapy for breast and prostate cancer was addressed. Interestingly, while anti-estrogen therapy is now important in breast cancer treatment, in the past, high doses of estrogen were used for certain breast cancers to induce apoptosis. The chapter notes that it is necessary to address the physiological type of breast cancer before commencing therapy, as some forms of the disease are hormone-therapy resistant. The chapter also goes into detail about anti-androgen therapy for prostate cancer, and this is well done, although analysis of quality-of-life issues would have strengthened the discussion of pharmacology-based treatment options for this disease.

#### 2.8 Special systems pharmacology

The dermatological and ocular pharmacology chapters have been greatly expanded compared to the 12^th^ edition of _GG_PBT. Five new agents (bentoquatam, coal tar, anthralin, brimonidine, and propranolol) were added to the “miscellaneous agents” section and one agent was removed (podophyllin) in the dermatological chapter compared to last edition. Both chapters have high quality colorful figures of pathways and tables with details such as structural class and efficacy for each agent. Additionally, there is a drug summary table at the end of the chapter with therapeutic uses, clinical pharmacology, and tips for each agent to compliment the text.

The ocular chapter provides excellent explanation of the medications’ mechanisms of action and describes which agent is preferred or which agent provides fewer side effects compared to an agent within the same class. For example, brimonidine is less likely to cause ocular allergy and for that reason, it is more commonly used. Although this information is presented throughout the chapter, it is not accessible in one place. An algorithm highlighting what is first-line and second-line treatment for glaucoma in the ocular chapter would have been beneficial to the reader.

When compared to Koda Kimble 10^th^ edition, ^29^ the dermatological and ocular chapters in _GG_PBT lack cases. It would have been helpful to have included case questions within the chapter, so the reader can easily see how the pharmacological information is applicable to patients in practice. In comparison to _Kat_BCP, ^23^ the _GG_PBT dermatology chapter reviews more medications. However, the medication section headlines are not as clearly defined and are presented in a less organized format. Within the sunscreen portion in the dermatological chapter, it might have been desirable to have more details about each agent. The chapters use only the generic name of medications. Since pharmacy students are expected to know both brand and generic names for the NAPLEX and in practice, it would have been appropriate to use both within the tables. In contrast, use of brand names would be less useful for medical students. Additional information from a student perspective may be found in the Supplemental Appendix.

## DISCUSSION

The quantitative and qualitative evaluation provided here is a substantial extension of prior reports.^1-8,12^ Evaluating a large (currently > 1,440 pages) textbook^1-7^ in just a few paragraphs, as is common, loses some nuance and may do a disservice to the breadth of coverage for an interdisciplinary and rapidly changing field. Overall, the editors and authors of _GG_PBT should be recognized for assuming such a large task as each new edition is a substantial and commendable achievement.

Approximately four out of every five _GG_PBT authors were male. Unfortunately, this finding is concurrent with a larger body of evidence.^9, 10^ For example, a widely used internal medicine textbook had slightly fewer (18.6%) female authors.^12^ The gender gap in _GG_PBT authorship showed some improvements relative to the 12^th^ edition where eight out of nine contributors were male.^8^ At this rate, _GG_PBT may achieve gender parity when the sixteenth edition is published in 2039. We are not suggesting that editors should recruit authors to achieve an arbitrary numerical representation. However, if the expertise exists, as is evident in some pharmacotherapy textbooks,^8,12^ and the pool of authors insufficiently reflects this diversity, then the policy that _GG_PBT selects authors would merit reconsideration.

Citations in _GG_PBT were significantly less recent than those found in both _Kat_BCP and _DiP_PAPA. Further, references were of similar age in _Ka_tBCP. The difference between sections with the most recent and oldest citations was only 2.7 years versus over twice as large for _DiP_PAPA (6.5) and _GG_BCP (6.7). Agents used for gastrointestinal disorders were most recent in _GG_PBT and _DiP_PAPA while drugs acting on the nervous system were the most dated in _GG_PBT and _Kat_BCP. Possibly, the neuropharmacology sections may change with further development of novel agents for Alzheimer’s^29^ and neuroendocrine pharmacotherapies for obesity.^30^

There is also substantial room for improvement in conflicts of interest transparency in _GG_PBT. With rare exceptions^21^, biomedical textbooks do not currently report on CoIs. Even in _Dip_PAPA, which discloses self-reported CoIs, four of the top ten highest compensated authors appeared to under-report their disclosures which could be due to a narrow reporting time frame, a narrow definition of “relevant” CoIs, or failing to provide the information to the editors. Editors may need to formalize responses to inaccurate disclosure including reductions to the honorarium or exclusion from future editions. Primary sources often employ the International Committee of Medical Journal Editors (ICMJE) ^31^ form for disclosure of potential CoIs where authors are instructed to be inclusive. More specifically, the instructions specify “You should disclose interactions with ANY entity that could be considered broadly relevant to the work. For example, if an article is about testing an epidermal growth factor receptor (EGFR) antagonist in lung cancer, one should report all associations with entities pursuing diagnostic or therapeutic strategies in cancer in general”. On the other hand, the ICMJE instructions might need to be adjusted to incorporate the extended time-frame of textbooks where the same authors commonly contribute to multiple editions over an extended period. Verification of self-reported disclosures with ProPublica’s Dollars for Docs (_PP_DD) and Medicaid Service’s Open Payments (_CMS_OP) would be a trivial task for publishing staff. Complete and accurate CoI disclosures will contribute to maintaining the highest degree of trust that content is consistent with the principles of evidence based medicine^20^ in subsequent editions.

Narrative evaluations by content experts revealed several strengths but also some potential for improvement. The neuropharmacology section missed an opportunity to at least begin to acquaint future health care providers that terms in wide-spread use like “mood stabilizers”, “antipsychotics” and “stimulants” are not informative and could be replaced by neuroscience-based nomenclature.^24^ There is also a need for more uniform avoidance of stigmatizing language.^27^ In general, the chapters on neoplasia are sound, and give a reasonably comprehensive overview on both traditional pharmacological therapies as well as some of the newer, more cutting-edge approaches. There are some weaknesses, however. A better understanding of the basic molecular biology would be helpful; in particular, this would strengthen discussion about the development of resistance to theory, as well as how signaling pathways can be modulated for therapeutic effect. Other areas of improvement would be an increased attention to chemoprevention, novel future approaches, and quality of life issues concerning side effects of treatment. More broadly, identification of which agents are not available in the United States at the time of publication would benefit multiple sections

Some caveats and limitations are noteworthy. First, the sex differences identified are consistent with a broader pattern in academic medicine.^8-10,12^ Future research should determine whether ethnic minorities are similarly under-represented as biomedical textbook authors. Second, the CoI databases do not report on non-financial CoIs or the financial relationships of non-physicians (i.e. half of _GG_PBT authors). Therefore, the three million received by authors from pharmaceutical companies is likely an underestimate. The presence of potential CoI, although appreciable, does not necessarily mean that content was impacted in any way. Third, the textbook citation differences in Figure 1A would be less pronounced if _GG_PBT were published slightly (one year) earlier. Therefore, the findings within each textbook (Figures 1B, 1C, 1D) may be more meaningful. On the other-hand, while the findings in Figure 1A were hypothesized *a priori*, the results in Figure 1B-D were not and may be susceptible to Type I errors. Fourth, although the narrative reviewers came from diverse fields and had substantial experience as educators, health care providers, and biomedical scientists, collectively, we are a far less distinguished group than the esteemed authors of _GG_PBT. There is some subjectivity underlying expert opinion and it is likely that others may have identified somewhat different strengths and limitations in _GG_PBT content.

## CONCLUSION

This report identifies many strengths of the 13^th^ edition of _GG_PBT but also some areas for improvement including greater diversity of authors, heightened transparency of conflict of interest disclosures, and improved recency of citations in select areas.

## Data Availability

The raw data in Figure 1 is included as a supplemental file.

https://openpaymentsdata.cms.gov/

## ACKNOWLEDGEMENTS AND DISCLOSURES

This endeavor was completed using software provided by the Husson University School of Pharmacy and the National Institute of Environmental Health Sciences (T32 ES007060-31A1). BJP is supported by the Center of Excellence, Health Resources Services Administration (D34HP31025) and Pfizer. DYK is currently employed with a biotechnology company. John Szarek, PhD provided feedback on an earlier version. This report is based on the analysis of the authors and does not reflect the opinions of our respective institutions.

## Figure Caption

**Supplementary Figure 1.**
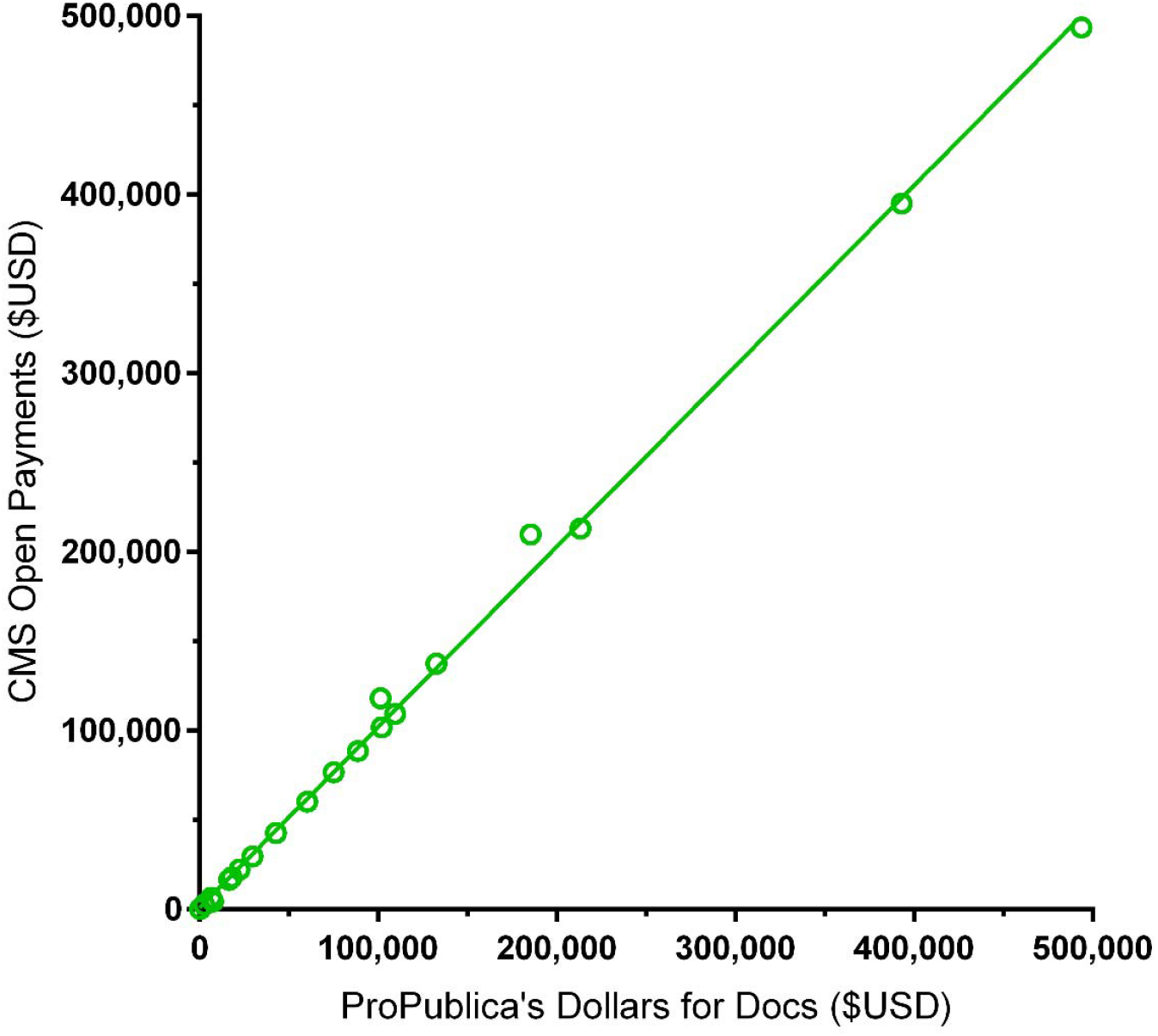
Scatterplot depicting the similarity in undisclosed potential conflicts of interest among Goodman and Gilman’s *The Pharmacological Basis of Therapeutics* (_GG_PBT) authors as reported by ProPublica’s Dollars for Docs (_PP_DD) and the Center for Medicare and Medicaid Service’s (CMS) Open Payments. R^2^ = .9978, *p* < .0001. Two authors (e.g. oncologist CI = $185,223 in DD vs $209,943 in OP) had more payments reported by Open Payments.

**Supplemental Table 1.** Comparison of self-reported and mandated reporting of conflicts of interest among the ten highest compensated authors of DiPiro’s Pharmacotherapy: A Pathophysiologic Approach, 2017. ^N^name not listed in the disclosures provided by Access Pharmacy 12/10/18.

| <u>Author: Chapter</u> | <u>Self-Report</u> | <u>ProPublica's Dollars for Docs 2013 to 2016</u> |
| --- | --- | --- |
| KD: Parkinson Disease | not reported<sup>n</sup> | \$729,695, \$349,614 for Parkinson Disease treatments (Apokyn: \$132,702, Azilect = \$111,326, Duopa: \$34,934, Deep Brain Stimulation: \$1,200) |
| JWW: Status Epilepticus | "none" | \$644,986, \$533,633 for anti-epileptic drugs (Fycompa: \$235,680, Oxtellar: \$78,942, Sabril: \$62,700, Onfi: \$58,630, Banzel: \$49,082, Qudexy: \$24,122, Aptiom: \$11,144, Trokendi: \$10,082, Vimpat: \$3,521) |
| ESR: Urinary Incontinence | "Consultant: Allergan, Astellas Pharma, and Ferring Pharmaceuticals" | \$227,510, \$131,600 for Urinary Incontinence treatments (Botox/Allergan: \$78,260, Myrbetriq/Astellas: \$25,584, Toviaz/Pfizer: \$10,815, Vesicare/Astellas: \$8,790, Interstim/Medtronic: \$8,151) |
| AM: Multiple Sclerosis | not reported<sup>n</sup> | \$99,634, \$69,907 for Multiple Sclerosis treatments (H.P. Acthar: \$16,034, Copaxone: 11,591, Plegrity: \$10,122, Lemtrada: \$8,698, Tysabari: \$8,630, Aubagio: \$6,086; Alemtuzumab: \$5,601, Bateseron: \$3,145) |
| SSCR: Gastroesophageal Reflux | not reported<sup>n</sup> | \$53,588, \$0 for Gastroesophageal Reflux drugs |
| DCH: Stroke | not reported<sup>n</sup> | \$14,762, \$125 for Thrombectomy device |
| DJL: Pulmonary Arterial Hypertension | "none" | \$13,573, \$10,817 for Pulmonary Arterial Hypertension treatments (Letaris: \$4,142 Orenitram, \$2,251, Tyvaso: 2,219, Remodulin: \$2,205) |
| JP: Pulmonary Function Testing | "none" | \$2,716, \$0 for Pulmonary Function Testing |
| MSH: Drug Induced Pulmonary Disease | "none" | \$2,045, \$0 for Drug Induced Pulmonary Disease |
| JMC: Colorectal Cancer | "none" | \$1,824, \$0 for Colorectal Cancer treatments |

## Supplementary Appendix A Evaluation of two master’s students

#1: The new edition of *Goodman and Gilman’s Pharmacological Basis of Therapeutics* (_GG_PBT) seeks to explain the concepts of pharmacology beginning with the basics and continuing through some of the practical applications used in many hospitals today. The organization of the text is superb and allows for easy comprehension. After the basics are covered, the physiological systems and their drug targets are neatly categorized so as to help the reader understand the interactions between the biological system, the underlying disease, and appropriate medication. After each of the tissue or system specific maladies are covered, the text delves into more broad illnesses and their treatments in the infectious disease and chemotherapy sections.

The writing in the text is clear and easy to understand. Each disease is well defined, and the text makes good use of diagrams and figures to explain the mechanisms behind the illness. After elucidating the cause of the problem, the text explains the mechanisms behind each potential treatment, and clearly summarizes their advantages and risks in a table. Any unusual or dangerous interactions of a drug are sufficiently explained, and the precautions to using such a medication are always described.

#2 The 13^th^ edition of *Goodman and Gilman’s Pharmacological Basis of Therapeutics* (_GG_PBT) made the learning and interpretation of pharmacology easier for the reader. The way the content is organized, the reader is better able to pick out important ideas from the support given on each topic. This provides a good outline for studying the material required by any specific course using this textbook.

The order of the text overall offers a lot of information at the beginning, and towards the end there are applications of the reader’s knowledge to the treatment of specific diseases and uses for some of the drugs. The text also provides an explanation for physiological processes before explaining the drugs that affect them, which allows the reader to have a better understanding of the significance and action of the drug. Each chapter is formulated so that the material is interesting, and difficult subjects are explained well so that the reader is not lost in the details. There is sufficient support given for the content, so the reader can apply what they are learning to other outside sources and ideas. Clear and legible figures and tables are placed throughout the text in order for the reader to visualize the content in a different form. Explaining the image well within the text provides a good flow that allows the student to understand the topic better. The drug summary tables that are provided are informational and concise which offers the reader a good summary resource. The antipsychotic molindone is listed as “not available in the US” in Table 16-2 but was listed in October 2018 as available^1^. An online textbook can be regularly updated.

Although the text does engage the reader, _GG_PBT does not provide sufficient critical thinking points throughout the text. There are no problem-solving opportunities for each topic, which limits the application and testing of material. The student would need to purchase yet another book for this.^2^

## Footnotes

1 https://dailymed.nlm.nih.gov/dailymed/drugInfo.cfm?setid=1275e92f-9573-4d0c-8e77-1c9ac47696d2

2 Rollins DE, et al. Workbook and casebook for Goodman & Gilman’s The Pharmacological Basis of Therapeutics, 2016, McGraw Hill.

## Notes

### Author Declarations

All relevant ethical guidelines have been followed and any necessary IRB and/or ethics committee approvals have been obtained.

Any clinical trials involved have been registered with an ICMJE-approved registry such as ClinicalTrials.gov and the trial ID is included in the manuscript.

